# Poverty Proofing health care: a qualitative study of barriers to accessing health care for low income families with children in Northern England

**DOI:** 10.1101/2023.10.04.23296541

**Authors:** Elaine Bidmead, Louise Hayes, Emma Leggott, Josephine Wildman, Judith Rankin, Luke Bramhall, Liz Todd, Laura Mazzoli-Smith

**Affiliations:** Institute of Health, University of Cumbria, Carlisle, Cumbria, United Kingdom; National Institute for Health and Care Research (NIHR) Applied Research Collaboration (ARC), North East and North Cumbria, Newcastle Upon Tyne, Tyne and Wear, United Kingdom; National Institute for Health and Care Research (NIHR) Research Development Service (RDS) North East and North Cumbria, Population Health Sciences Institute, Newcastle University, Newcastle upon Tyne, Tyne and Wear, United Kingdom; Children North East, Newcastle upon Tyne, Tyne and Wear, United Kingdom; Population Health Sciences Institute, Newcastle University, Newcastle upon Tyne, Tyne and Wear, United Kingdom; School of Education, Communication and Language Sciences, Newcastle University, Newcastle upon Tyne, United Kingdom; School of Education, Durham University, Durham, County Durham, United Kingdom

## Abstract

Poverty impacts negatively on children’s health and future life chances. Access to the UK’s National Health Service (NHS) is based on clinical need rather than the ability to pay but horizontal inequities in access exist. Children North East, a charity supporting children experiencing poverty, wanted to develop a Poverty Proofing© Health tool to help NHS services reduce the impacts of poverty on access. This study aimed to understand barriers to healthcare access faced by families living on low incomes to support development of the tool. Twenty parents and seven Voluntary Community Social Enterprise sector staff participated in qualitative interviews or focus groups. Data were analysed thematically, and three main themes were identified as impacting access to health care: hidden costs, securing appointments and developing relationships with health care providers. We conclude that low-income families experience both financial and other barriers to accessing NHS health care and that these barriers are exacerbated for low income families living in rural areas.

## 1. Introduction

Poverty is a major determinant of health and life opportunities that impacts negatively on children’s futures. Poverty occurs when people’s resources are well below their minimum needs [1]. A recent UK report shows that ‘children born into the poorest fifth of families in the UK are almost 13 times more likely to experience poor health and educational outcomes by the age of 17 years’ [2]. Children experiencing poverty are at greater risk of becoming overweight, developing asthma and having tooth decay, as well as performing poorly at school [3]. Poverty also increases the risk of poor mental health [4] and early adulthood mortality [5,6]. Children and young people in the UK report that poverty has a negative effect on their wellbeing and causes feelings of exclusion, shame and unfairness [7].

In 2021-2022, 29% of UK children lived in poverty [8]. In June 2023 the figure was substantially higher (35%) in the North East of England [9]. Poverty is a significant cause of the UK North-South divide in children’s life chances as it limits access to the circumstances that help to ensure good health [10].

Children North East (CNE) is a charitable organisation founded to support children living in poverty. In 2011, children working with CNE identified an end to discrimination in school as their main priority. In response, CNE, with support from the North East Child Poverty Commission, developed ’Poverty Proofing© the school day’. This is an audit and action-plan development tool for schools which aims to remove poverty-related barriers to learning [11]. Following several successful implementations, the North East and North Cumbria Child Health and Wellbeing Network expressed an interest in developing Poverty Proofing approaches for health care settings.

Access to the UK’s National Health Service (NHS) is based on clinical need rather than an individual’s ability to pay. It is free at the point of delivery irrespective of financial circumstances. However, ’horizontal inequities’ exist in health care utilisation [12]. While poorer health in poorer people results in them consuming more health care (at every age), richer people tend to access health care at an earlier stage and consume more preventive and specialist care [12, 13].

This study aimed to identify barriers to health care access among families with children living on low incomes (commonly defined in the UK as below 60% of the median income [14]) with the objective of designing an action-plan toolkit for Poverty Proofing a range of health care settings. The work was conducted during a period when the NHS was experiencing service pressures due to increased demand from COVID-19, workforce issues, and the impacts of austerity. Comments during data collection made clear that these pressures had created a situation in which access to health services (such as appointments with dentists, General Practitioners (GPs) and hospitals) was challenging for many people, irrespective of financial circumstance. In this paper we focus on issues impacting access to health care that are amplified by low-income. The results presented informed the development of a Poverty Proofing audit tool.

## 2. Material and methods

The COnsolidated criteria for Reporting Qualitative research (COREQ) [15] were used to guide our research design; our COREQ checklist is attached as appendix A.

### 2.1 Sample and Recruitment

Data collection was undertaken with parents on low-incomes and with professionals working in the Voluntary, Community and Social Enterprise (VCSE) sector who support people on low-incomes.

Parent participants were purposively recruited via VCSE organisations in the North East and North Cumbria. VCSE organisations were supplied with study information sheets which they shared with service users they identified as experiencing low-income (number unknown). Participants were offered a £20 gift voucher to thank them for their contribution. Twenty parents volunteered; none were known previously to the researchers.

Three VCSE organisations working in health and wellbeing were contacted by email, supplied with a study information sheet, and invited to contribute to the research. Organisations were offered compensation of £20 per person participating. Eight staff members volunteered to participate. All were based in North Cumbria and two were known to EB previously.

No participants dropped out. The number of parents/VCSE staff choosing not to participate is unknown.

### 2.2 Ethics

Ethical approval for the study was granted by Newcastle University Research Ethics Committee (Reference: 2236/15258 Date: 23/11/2021).

All participants were provided with written and verbal information about the study and invited to ask questions prior to participating; all provided written informed consent before taking part.

### 2.3 Theory, design and setting

This study employed a critical realist, contextualist approach to gather and understand participants’ situated experiences of accessing NHS health services whilst living on low incomes.

A topic guide was devised to structure and facilitate in-depth one-to-one interviews and focus groups; it was piloted with two parents and no amendments were made. The topic guide prompted participants to think about different aspects of health care, including getting appointments, accessing appointments, emergency situations, and staff attitudes; it also allowed participants to introduce issues significant to them.

The research was undertaken by EB and LH. Both are female, mid-career researchers (educated to PhD level) trained and experienced in qualitative data collection methods and analysis. Researchers shared with participants their names, job roles, qualifications, and reasons for doing the research.

One round of data collection was undertaken. Interviews with parents were conducted via online video using Microsoft Teams or Zoom, six by LH and one by EB. Interviews lasted between 15 and 60 minutes and were recorded and fully transcribed. Three parent focus groups were conducted in community venues and facilitated in person by LH. Parent focus groups lasted between 40 and 60 minutes; two were audio-recorded and fully transcribed; notes were taken in the third. The interview and two focus groups with VCSE staff were conducted by EB via online video using Microsoft Teams, they lasted between 60 and 80 minutes and were recorded and fully transcribed. No others were present during data collection besides the researchers and participants. Transcripts were not shared with participants.

### 2.4 Timescale

Data were collected between December 2021 and March 2022.

### 2.5 Analysis

Anonymised transcripts were analysed using ‘reflexive thematic analysis’ [16]. Following familiarisation with the data, a sample of six transcripts were independently coded by two researchers (EB and LH), inductive codes emerged from the data; deductive codes aligned with the topic guide. The two researchers then met to discuss codes and construct initial themes. Having agreed the coding frame all transcripts were coded by one researcher (EB) using NVIVO. Following coding, initial themes and coded data were discussed at team meetings and the main themes were agreed. EB and LH drafted the paper except for the discussion which was drafted by EL. The paper was reviewed by JR then commented on and agreed by the study team and finalised.

## 3. Results

Semi-structured interviews were conducted with six parents (all mothers) aged between 20-60 years, four of whom were lone parents. Thirteen parents (12 mothers and one father), aged 20-40 years, participated in one of three focus group discussions. Seven staff took part in focus group discussions and one in a semi-structured interview; all were female and aged over 30 years. The results presented below include quotations to provide rich and faithful accounts; each transcript was given an identity code to indicate whether data came from a parent interview (ParentInt), parent focus group (ParentsFG), VCSE interview (VCSEInt) or VCSE focus group (VCSEFG).

Contributors commented on the challenges of living in poverty which we discuss first. Next, we present thematic findings. We identified three main themes: hidden costs, securing appointments and developing relationships with health care providers. We then discuss contributors’ awareness of sources of financial assistance for accessing health care followed by contributors’ suggestions for improvements.

### 3.1 Living on a low income

Some parents shared the challenges of living with financial hardship. It was typically described as hard, stressful, embarrassing and stigmatising:

“It’s a big struggle on a on a day-to-day basis It’s like this week it’s my, my poor week as I call it, I get like £63 today to last me a week and out of that I’ve got my bills to pay, my food to pay it’s just impossible to live on.” (ParentInt1)

Participants in ParentsFG4 spoke about the embarrassment felt at having to ask for assistance due to financial hardship. ParentInt2 shared the stigmatising effects of poverty: “stigma about people claiming benefits, and ‘Oh, you’re useless,’ and, ‘You’re timewasters and scroungers.’” They stressed that they had always worked until their child was born with significant health needs and explained that they were “quite proud”; being asked to prove they were in need was highly embarrassing to them, for example:

“You either have to print off a statement, which to me is embarrassing, because it’s got every bit of how they break down how much you’re going to get paid the other thing that I find quite embarrassing is, if, for example, my son’s admitted into hospital, they’ll ask me about what our situation is. Do I work or don’t I work? And do we have social worker? And I feel that’s quite probing I think there’s a lot of things that could be improved on, to make you feel not as scummy about the way you are.” (ParentInt2)

VCSE contributors described what living in poverty meant to them. Descriptions highlighted both absolute poverty (reliance on food and clothing banks) and relative poverty (having to do without luxuries, holidays, family days out etc.). Living in financial hardship was thought to impact people’s long-term health and resilience due to poor diet and limited social engagement (VCSEFG1). VCSE contributors also emphasised the stress caused by poverty and the complex, layering and cumulative impacts it has on families, which can become overwhelming.

“Poverty … It’s an overwhelm with lots of different factors that eat you away, until you feel like you have few options.” (VCSEInt1)

“It’s bad enough if you have a child with a physical need … If you add on the fact that if there’s poverty and you’ve got a battle with the health system … you know it’s all about those layers.” (VCSEFG2)

Like parents, VCSE contributors highlighted people’s embarrassment about their financial situations because “families often feel judged” (VCSEFG2). Consequently, some attempt to hide or disguise the effects poverty has on their lives.

VCSEFG2 pointed to situations where the impacts of poverty may be misconstrued and understood instead as neglect. For example, one participant highlighted the practice of setting targets for parents during Child Protection or Child in Need Plan meetings, such as attending health appointments. They noted “if you weren’t actually able to access the appointment because of difficulties getting there” then that may be seen as the parent not complying:

“Some families will go down the I can’t afford it. Others won’t say that because they’ll be fearful of ‘Well, if you can’t afford to go to health … what’s happening there’ and they will come over as disengaged and difficult when actually they can’t, they cannot, actually manage to get there because of finance.” (VCSEFG2)

### 3.2 Hidden Costs – financial barriers to accessing health care

#### 3.2.1 Transport

Transport costs emerged as a major barrier to accessing health care for low-income families. Most could access GPs locally and so could attend relatively easily, but attending hospital appointments often involved longer journeys and several buses if undertaken by public transport, or excessive parking fees if undertaken by car.

Contributors indicated numerous times that the costs of transport to appointments was a concern to them and these costs increased exponentially for parents with multiple appointments, living in remote/rural areas, or having to travel long distances to access secondary care. There were also significant time implications for parents living in remote areas without access to a vehicle, one parent from a rural village elaborated:

“It’s about 25, 26 mile away so like I say to get to [large town], you’ve either got to get a taxi … to [small town] and then a train … to [large town] … or get a bus which takes about 45 minutes. The train takes about 10 minutes, but they don’t run as often … For a taxi into [small town] its £15 each way … The train I think it’s about £7 or £8 return now … for both of us, you’re looking, by the time you get taxis and that, you’re looking at 50 odd pound before you even get to the appointment […] So, you have to cancel appointments or work around it.” (ParentInt1)

In rural areas, people may be required to access secondary care at sites across the region, irrespective of where they live. Where people are referred is determined by the availability of appointments. However, people on low incomes face the dilemma of covering significant costs or delaying their health care:

“Where health authorities are trying to squeeze you in for an appointment and not make you wait, they will offer you say [hospital name] for example … for some families, that’s easy enough [but] often it’s a choice of getting yourself over to another hospital … taking the day off work or waiting maybe a month or two, you know, for your child to be seen, which is a really difficult decision to make […] they say you aren’t forced to go there, you can wait so that while you’re not denied health care it will be delayed, not through your lack of engagement, not through you not putting your health or your children’s needs first, it’s because of the affordability of the offer of what your appointment looks like.” (VCSEFG2)

When it comes to accessing tertiary care, such challenges are exacerbated. VCSEFG1 commented upon the number of people in North Cumbria having to travel “out of county” for treatment. For example, for specialist children’s services, “Newcastle’s the closest or … it’s Manchester” (VCSEInt1). This entails expensive and time-consuming travel and potentially lost earnings if time off work is necessary. One parent in rural North Cumbria highlighted the journeys involved in accessing care for their child ‘out of county’:

“I went to see [consultant], so of course that incurred money for going across to [hospital in Newcastle], and then he asked for an MRI scan, which [hospital in North Cumbria] did, and then he sent me an appointment to have an MRI scan done with contrast, but at [another hospital in Newcastle].” (ParentInt2)

This means some people simply cannot afford to attend hospital appointments, a situation that is compounded where multiple appointments are involved.

#### 3.2.2 Subsistence during hospital attendance

Paying for food and drinks during hospital attendance emerged as a significant challenge for parents who could not afford the costs involved; this was an issue when children were outpatients and inpatients. Parents highlighted the limited options within hospitals to buy food, and stressed the expense of hospital food outlets in the North East and North Cumbria:

“Hospital food is not affordable … I just had to find the money. My mam would help us out a lot. My daughter’s grandad would help. But it is still too much to pay back.” (ParentInt5)

“Even getting a cuppa, it’s like a fiver in the hospital.” (ParentsFG5)

Costs increased when children were inpatients. It was reported that no food is given to parents staying with a child, no matter their age. One parent had experienced extended hospital stays when their baby was born with health complications. They stayed in charitable accommodation at hospitals; these provided cooking and laundry facilities. However, they did not regularly use these due to financial and time pressures which then impacted their ability to eat healthily; they reported missing meals and not looking after themselves properly. ParentInt6 noted: “The food in the hospital, it’s not conducive to low cost” and reported “living on Subway sandwiches and Pot Noodles but I can’t see it being nutritionally balanced” and that “I try to pack snacks for both of us”.

Whilst some preparedness may be expected for planned admissions, this is not possible with emergency admissions which were reported to trigger even more expenditure. VCSEInt1 highlighted the realities of emergency admissions for parents:

“As we know from our stats, that’s likely to be an ambulance admission. So, they’re less likely to have what they need with them. They’re less likely to have the people that they need with them […] you’d be lucky if you find a vending machine. You’d be lucky if you’ve got the right change on you for the vending machine and you’d be lucky if the vending machine’s got anything in it other than tea or coffee. I think you might find that a kind nurse might offer you a cup of tea and a piece of toast, if they’re not rushed off their feet.”

This scenario was confirmed by parents, for example:

“I didn’t have a budget for when he got rushed into hospital, because nobody knew it was going to happen. So, then I was borrowing money and things like that, just to travel over there and back. (ParentInt1)

On a positive note, two parents mentioned specific nurses or wards being helpful in terms of providing food and drinks, but this was dependent on the actions of individual staff members rather than hospital policy.

#### 3.2.3 Discharge from hospital

Participants highlighted the costs associated with discharge from hospital following an admission by ambulance. This is a particular issue when the emergency department is a significant distance from home and if discharge occurs when public transport is not operating. Hospital discharge “could be any time of the day; it could be at any time of the night” (VCSEFG2) and “Your only option for getting home without any transport would be a taxi” (VCSEInt1). Parent5 described attending emergency departments several times; each visit requiring a taxi. Sometimes their child would be admitted, but at others they would give medication and discharge them, “so that would be another £30 taxi fare back home” (ParentInt5).

#### 3.2.4 Parking costs

For parents who owned a vehicle parking charges were a concern; they were said to be around “£2 or £3 an hour” which “obviously you don’t have” (ParentFG5). This view was repeated by VCSEFG1 where it was mentioned that “parking is always an issue because of the extra costs parking brings” which caused stress “because you’d never know how long” you will need. ParentInt2, whose child was in hospital for several weeks, reported high parking charges when their partner drove from North Cumbria to Newcastle at weekends. After a time, somebody mentioned cheaper parking that could be used but this involved using a shuttle bus to the hospital, which took up time and was not available at weekends.

#### 3.2.5 Impacts on income

Some contributors spoke about lost income due to attending appointments with children. ParentInt2 reported losing pay from having to miss work when their child was ill because they did not “qualify for sick pay”. A contributor in VCSEFG2 highlighted that some families cannot take time off work because it will affect social security payments.

For others, having an ill child had resulted in them losing employment. Contributors in VCSEFG2 talked about several parents who gave up employment because they found working whilst caring for an ill child impossible and decided it was “too much to be able to juggle all these things and meet my child’s needs”. Another added that “the way that health services shape themselves impacts” because people must choose between “working or putting children first”; they highlighted “the amount of negotiation you have to do to try and get not every single health appointment in the middle of the day. You know, it’s, it’s impossible” (VCSEFG2).

### 3.3 Securing Appointments

#### 3.3.1 Digital first

The issue of digital access to health services was raised on several occasions, as was the expectation that everyone has a smart phone or internet access with unlimited calls/data. Parents questioned if people on low incomes did. For example, whilst some participants at ParentsFG7 noted the easiest way to get a GP appointment was via an app, one shared that they did not have a smart phone nor access to a computer.

VCSE contributors referred to a digital divide wherein “you are less likely to have a laptop if you don’t have the money” (VCSEFG1). COVID-19 was seen to have accelerated the move to digital first services, which many people have adapted to over time, but “other people haven’t been able to just because they can’t afford it” (VCSEFG1). A participant in VCSEFG2 pointed to the requirement to have a mobile phone to receive a code that enabled one to access lateral flow tests for COVID-19. VCSEInt1 noted that:

“We absolutely know that the majority of people [experiencing financial hardship] need face to face engagement because they need to build up trust with their caregiver and … we do know that a lot of people will avoid digital connection and will only go for face-to-face connection because that’s the way that they work. That’s the way they’ve always operated.” (VCSEInt1)

#### 3.3.2 GP Appointments

Whilst not directly related to family poverty, almost all participants described difficulties in securing GP appointments, for example: “not being able to get appointments when you need them waiting weeks and weeks, and weeks” (ParentsFG3). Others reiterated this experience; one “waited two months” for a GP appointment regarding “a water infection” by which time “it was a kidney infection” (ParentsFG3). VCSE contributors also highlighted the difficulties experienced by service users in getting appointments due to “massive waitlists, wait times” (VCSEInt1).

Having to telephone the surgery for appointments was highlighted as problematic, participants described waiting on hold for long durations or having to call multiple times:

“I was in the queue for half an hour or 45 minutes. And then she said, ‘It’s really busy, I can’t give you an appointment, you can try tomorrow.” (ParentInt4)

“You’re having to ring 30 to 90 times to get through … I think the record is 124 to get an appointment one morning.” (ParentInt6)

Specific issues with telephone consultations were also reported. Contributors resented having to wait in for doctors to telephone as “they won’t give you a time for them ringing you back … you’ve got to be free all day. If you miss that day, then you’ve really had it” (ParentsFG5). ParentInt1 reported similarly and commented “It stops me from ringing the doctors more times than enough to be fair” (ParentInt1).

Two important factors emerged for low-income families alongside these issues. First was the cost of being on hold for extended durations. VCSE contributors pointed to an assumption that all patients have access to free calls on their home or mobile phones and have data for online access, which is not the case for families facing financial hardship, who are more likely to be on a basic landline tariffs or pay as you go and have limited data on their mobile phone.

“If it’s ringing, it’s free and people just assume that people don’t want that, and they don’t want to be ringing, they want to be in a queue. But … that starts the pennies ticking for people who have to pay for their calls.” (VCSEFG2)

The second issue to emerge was that these difficulties discouraged people from trying to obtain appointments, potentially exacerbating conditions, with some choosing to access Accident and Emergency Departments instead:

“They’re not even bothering … because they can’t get through to them. So, they won’t even bother ringing, they won’t ring for the children, they won’t ring for their husbands and won’t ring for themselves anymore and they will go, if it’s an emergency, they would go to the hospital. But apart from that they’ve not even tried, they’ve just stopped ringing because they don’t, they just don’t see the point anymore.” (VCSEFG1)

“The system seems quite complex to navigate, sometimes you’ve got to ring up and then listen to something … some of my families find that really difficult, and they find it so frustrating. I’ve actually got a family who are more likely to ring, do a 999, you know, call a blue light, than actually, you know, ring through for … a GP appointment.” (VCSEFG2)

Contributors in VCSEFG1 also highlighted issues in getting appointments for those working more than one job, non-standard hours or with inflexible employment terms who might not be able to take time off work, and for whom time off may result in loss of income. Consequently, parents had not attended appointments “because they just couldn’t get time off work” (VCSEFG1).

#### 3.3.3 Dental Appointments

The difficulties of accessing NHS dental care are a national problem that has been highlighted extensively in national media in recent months. Indeed, many people have little option but to access dental treatment privately “but that’s not the case for people that don’t have the money to do that” (VCSEFG1). However, even NHS treatment is expensive to those on low incomes. ParentInt5 explained that charges might prevent them from seeking treatment: “if I had to pay for that extra treatment, I wouldn’t get it done” (ParentInt5).

Moreover, participants in ParentsFG7 reported routinely waiting 12-18 months for appointments for their children, during which time their oral health had deteriorated and children were in pain. This scenario was repeated in ParentsFG5, for example: “Trying to get an appointment at the dentist is like a needle in a haystack. You can’t get one”; “they said I couldn’t get an appointment until next year”.

One VCSE contributor in VCSEFG1 suggested that poor dental health is once more becoming a marker of poverty.

#### 3.3.4 Navigating and negotiating appointments

VCSE contributors commented on the difficulties some families face in navigating and negotiating health appointments. The health care system was described as a “minefield” involving complicated referral systems and criteria, and what often feels like arbitrary decision making. The system was said to be challenging to negotiate even for those with knowledge of it; for families without such knowledge, it can be debilitating and exhausting, as well as expensive and resource intensive due to having to make multiple telephone calls and searches for information:

“It feels like they’re having to jump through the hoops … they might be feeling are we at the bottom of the pile? Or is it just because of COVID? Or is it just that, you know, our needs are being ignored” (VCSEFG2).

VCSEInt1 highlighted people’s inability or reluctance to advocate for their health care and that of their families as well as poor awareness of what is available to them “so they’re not even accessing those support and services. You know they’re not asking for the referral.” VCSEInt1 ventured that few of us are “good at asking for help when we think we should” and most of us “leave it too late,” but where there are poverty issues “you leave it even later”.

#### 3.3.5 Appointment times

Parents commented on the difficulty of getting children to appointments due to having no control over appointment times. Appointment times determined by health care providers often clashed with other responsibilities, such as taking other children to school and work. Some preferred appointments outside the school day to avoid missed education for children, but others preferred appointments during school times so they did not have to take the whole family with them.

ParentInt6, whose child was on a child protection plan, spoke about how hospital appointments could sometimes clash with requirements of the plan, but hospital appointments were not easy to change, “a lot of them are … an afternoon clinic once a month or you have to go to this hospital 16 miles down the road”. VCSEInt1 also commented on the difficulty of altering hospital appointment times, even where long journeys are involved, “I think you’re entirely at the mercy of what comes through on the letter.”

#### 3.3.6 Childcare

Parents talked about challenges in finding suitable childcare so that they or their other children could attend appointments. Participants in ParentsFG3 talked about the possibility of using breakfast or after school clubs, but this was more expense. Moreover, several parents reported not having anyone readily available to provide childcare. This situation was recognised by VCSE contributors who reported that childcare impacted access to health care for parents and children alike. Both VCSEFG1 and VCSEFG2 referred to whole families presenting at accident and emergency departments because they had no one to leave children with.

### 3.4 Relationships with health care providers

Some parents commented on difficulties they had experienced in forming trusting relationships with health care staff. Not receiving care consistently from the same health care professional and having to keep “explaining yourself over and over” was a particular issue for parents in ParentsFG7. Parents offered little comment on health care providers’ responses to poverty beyond recognising and valuing moments where health care staff went out of their way to be helpful – for example, in the provision of food and drinks.

All VCSE contributors were supportive towards health care staff and acknowledged the pressures facing the NHS, for example: most people’s experiences of “health professionals have been positive; most people go into the job for the right reasons and are kind, caring and helpful” (VCSEInt1). Nonetheless, VCSE contributors held concerns over how health care providers respond to poverty: “organizationally there are obviously some issues” (VCSEFG2).

Contributors in VCSEFG2 contemplated whether clinical staff were trained or supported to think explicitly about poverty; in their experience, where solutions to poverty issues had been presented it had been “individuals who’ve done that as opposed to a systemic approach”. VCSEInt1 pointed to some health care staff that are “authoritative; don’t really listen” which they felt was potentially related to “training issues and recruitment issues in medical school” in that so many “medical students are from private school” and that “GPs tend not to do their placements in deprived areas” (VCSEInt1).

A contributor in VCSEFG2 pointed to an expectation within health services that patients can comply with available provision and when they cannot it is seen as a problem with the patient rather than the system. Another provided an example of when a single parent with four children did not take one of them to an “urgent appointment” because they had no childcare for the others. Whilst the parent had been supported to rearrange the appointment their decision prompted involvement from other services:

“So, [they] felt judged that, [they were not] able to get to that particular appointment at the correct time [because they] had made that decision that, actually, I can’t leave my children alone, and I’m going to have to stay with them.” (VCSEFG2)

VCSE contributors also pointed to issues with communications between patients and health care providers; they perceived that access to health care information was largely determined by an individual’s ability to navigate and communicate with the system, which was particularly challenging to people with poor literacy, health literacy and digital access/skills. VCSEInt1 highlighted the lack of services to support people who are “overwhelmed or just don’t quite understand or can’t cope”.

### 3.5 Awareness of financial assistance for health-related costs

There was a general lack of awareness about sources of financial assistance for health-related costs. The provision of such information appeared arbitrary, with some people gaining awareness by good fortune and others never becoming aware of it. Participants in ParentsFG7 commented about only finding out about support services and financial help by chance. ParentInt1 had “no idea about anything” and could not recall it ever having been mentioned. However, the idea of applying for assistance appeared burdensome: “If you’ve got to think about support on top of that to get to doctors and stuff like that, it’s another thing you’re having to constantly think about” (ParentInt1).

In contrast, ParentInt6 was aware that hospital transport might be available “but you have to be ready two hours before and you could be waiting up to two hours after”, which is not ideal when taking children. However, they reported that “if you ask about the transport, they can be quite shifty with you, especially if you’re young we need to keep it for the older generation, as I’ve been told” (ParentInt6).

Participants in ParentFG5 described how on many occasions they found out that they were eligible to financial support with attending healthcare services “only by chance”.

VCSE informants held little awareness of how to access financial support for health-related costs. VCSEFG1 pointed to “the amount of time it takes to look for the resource and support you can access” and commented that people “just don’t have the time to do that because you’re too busy trying to live your life; trying to work”.

### 3.6 Suggested improvements

Several suggestions for how to improve access to health care for families living on low incomes were made.

#### 3.6.1 Financial and practical assistance

ParentInt2 commented on awareness of help with financial costs for health care. They stated that had they known they were entitled to help due to low-income then they would have claimed but suggested that although they had read about assistance “in one of the leaflets, for one of the hospitals”, they were not sure whether they were entitled because “it doesn’t say how, it doesn’t say in what circumstances”. Therefore, ParentInt2 felt that information on help with health care costs should be included in letters sent out by health care providers and Universal Credit, something that says, “If you’re on this benefit, you can be entitled to this.”

Other parents suggested ways to help with subsistence. There were many comments about how hard and humiliating it could be to ask for financial support or for basic necessities to be provided when parents had to stay in hospital accommodation. Participants in ParentFG3 discussed being “embarrassed to ask” for things that would help but felt this would be easier if things were offered; they thought services should “offer, and overly offer.” ParentInt6 suggested providing vouchers to use at food outlets.

VCSE contributors all wanted increased awareness of financial support for health care costs. Contributors in VCSEFG1 identified knowledge about financial assistance to be a “big problem”. VCSEInt1 believed that at the very least, people should be made aware of available financial support, but also about any flexibility there may be around appointment times which would make attendance cheaper and more achievable. VCSEFG2 suggested awareness of assistance was so low, even amongst VCSE staff, that research was needed to clarify “what support is available” and for it to become “habit that health providers include that in their letters”. Both VCSEFG1 and VCSEFG2 suggested having leaflets and posters in areas of high footfall, schools, GP surgeries and all health care settings. Another contributor, in VCSEFG2, mentioned having seen posters on the doors of supermarket toilets which said “if you struggle to pay for sanitary products just go to customer service and ask for Sandy or something like that”. The group believed something similar would work in health care settings.

#### 3.6.2 System changes within health care

VCSEInt1 wanted to see more flexibility around appointments times:

“Because at the moment we just feel lucky if you get an appointment … it’s such a battle. But … if you’ve got a long journey, and especially if you’ve got other caring responsibilities or employment responsibilities, the more flexibility the better.” (VCSEInt1)

VCSEFG2 felt that patients should be able to “interact in the way that the doctor surgery wants you to at no cost.” They also wanted pre-bookable GP appointments made available to families, which they thought important for planning transport, especially for fitting appointments around the school day and other children’s care. Furthermore, for parents whose children have health conditions, getting GP appointments can feel like a constant trial so pre-bookable appointments would ease their stress: “you know they’re going to be going to the doctors in a month’s time. So why not?”

VCSEFG1 believed improving access and care for low-income families required “a whole cultural system shift”. VCSEInt1 highlighted the social and cultural (class) differences between senior health care professionals (i.e. GPs and consultants) and the communities they serve; they wanted medical students to experience more placements in deprived areas. In a similar vein, VCSEFG1 called for improved training for medical staff: “raising awareness about all these different kind of groups that don’t get access and why they don’t get access.” VCSEFG2 felt similarly, “training is really important … specifically about poverty and how that affects families.” VCSEFG2 admitted to having little awareness of how much training in poverty issues different health care professionals receive and acknowledged that some professionals are “seeing those issues all the time and [are] really aware of that”. Nevertheless, they felt training and awareness to be very important.

## 4. Discussion

The aim of this study was to identify barriers to health care access among families living on low incomes with the objective of designing an action-plan toolkit for poverty proofing health care settings used by children and their families. The following discussion summarises how the study has influenced the approach taken by CNE.

As a consultative model Poverty Proofing© Health (PPH) is grounded in the same ethos, principles and methodology as Poverty Proofing© the school day (table 1). However, the audit process for PPH has a wider training remit and a consultation phase that extends beyond the voice of the child to include a diverse range of people. This research provided evidence to strengthen CNE’s existing audit approach to PPH, especially in the areas of staff training and suggested improvements, and the themes identified resonated with those CNE see in practice.

**Table 1.** Ethical principles of Poverty Proofing©.

|  |  |
| --- | --- |
| **Voice** | To achieve real social change it is imperative that the voice<br>of those affected by poverty is central to understanding<br>and overcoming the barriers faced |
| **Place** | Alongside hearing from people, the context of the<br>community and place needs to be understood |
| **Structural inequalities** | Structural changes at an organisational level can be<br>addressed or at least alleviated by eliminating the barriers<br>that those in poverty face |

Following the research phase, CNE completed several PPH audits across different health care settings (including GP Surgeries, Maternity services, Outpatients, Paediatric Diabetes, Palliative Care, Sexual Health and Speech and Language). Whilst the terminology differed across settings there were clear similarities between the thematic areas emerging in the audits and those identified in the research (see table 2). This developed our thinking beyond an action-plan toolkit and led us to develop a ‘Common Themes Framework’. The framework presents CNE and the PPH audit process with a systematic, intelligence-led approach that underpins and sits across each of the five phases of the PPH audit process. How this translates into practice is explored next.

**Table 2.**
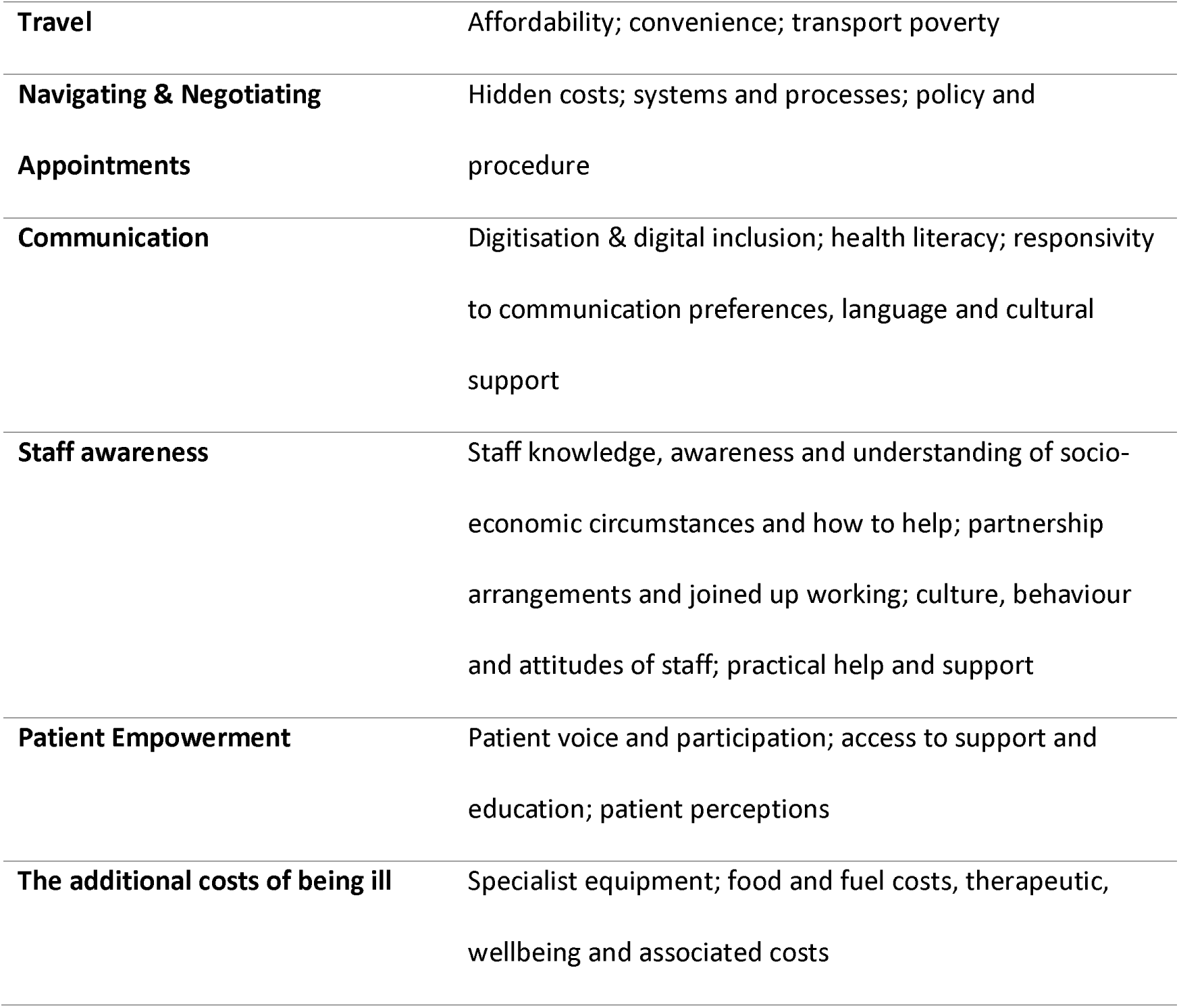
Thematic Areas in Poverty Proofing© Health.

### 5.1 The Development of a Common Themes Framework

A PPH audit has five phases, these are: (i) training and initial consultation with health care staff; (ii) scoping of the setting and how it works in practice; (iii) patient and community consultation; (iv) comprehensive report with recommendations; (v) review. In this section, we explain each phase, show how the common themes and suggested improvements that emerged in the research were integrated, and the impact this had.

#### 5.1.1 Training

Research participants felt health care staff would benefit from training, specifically about poverty and how it affects families. Whilst PPH always contained training, this phase has been revised to embed the common themes with the aim of raising awareness amongst health care staff of the barriers people face in accessing health care (i.e., travel, subsistence, accessing appointments, navigating the health system and relationships with health care providers). Facilitated consultation at the end of training sessions allows CNE coordinators to frame the consultation around these common themes and to draw out how staff experience the challenges of poverty in their work. This is essential to co-production, building relationships and understanding both structural barriers and the challenges staff face in delivering poverty informed services.

#### 5.1.2 Scoping

A set of prompts, relating to the research themes, have been designed and implemented to ensure PPH coordinators are informed and consistent in observations, conversations and paper-based enquiries whilst scoping and orienting the work in relation to each setting. Our scoping now includes gaining an awareness of the health care setting’s policies, processes and procedures that are most likely to impact families on low incomes (such as discharge policies and travel reimbursement schemes) and unpicking the mechanisms for signposting to support services, appointment booking systems and so on. This activity helps to discern between what is fact, what is policy and what is perception in both staff and members of the public.

#### 5.1.3 Consultations

The ‘Common Themes Framework’ also feeds into the way questions are posed during community consultations to emphasise those issues amplified by living on a low income and navigate away from questions that focus on challenges relating to those wider NHS pressures that most people experience, irrespective of income. The questions are designed to ensure that all common themes are addressed in a systematic way, whilst also allowing space for new themes and underdeveloped areas to emerge.

#### 5.1.4 Feedback & Reporting

A key feature of feedback and reporting is making recommendations and suggested improvements based on the findings from the PPH audit. CNE use a strengths-based approach that specifically seeks out what is working well in each setting and how it supports people living with poverty, as well as understanding the challenges and barriers to doing this. The improvements suggested by research participants are now included in a growing bank of recommendations that are drawn on when feeding back to organisations. PPH is delivered at place and the breadth and geography of settings means recommendations cannot be standardised or generalizable. However, having a bank of recommendations helps with efficacy and consistency where findings are similar. For example, travel is a topic that arises frequently. Therefore, understanding that the NHS low-income scheme includes travel reimbursement and ensuring the process for this is shared clearly and personally is one practical example of how health care staff can help close the travel inequality gap. Another is around asking about finances, universally, as part of patient care as a way of opening up the conversation so the right support can be identified. If this question is not asked health care staff can be unaware of patients’ financial difficulties and this can lead to assumptions, unconscious bias and missed opportunities to provide support.

#### 5.1.5 Impact and Monitoring

The ‘Common Themes Framework’ is reflexive in its nature and there is space within it to respond and grow to accommodate new knowledge and themes as they emerge. Structuring each audit around common themes means findings can be quantified and analysed by locality, theme and setting type. This then contributes to a broader framework of monitoring impact at scale, providing insight into common themes that contribute to health inequality in a way that can be pinpointed and articulated to policy and decision makers.

## 5. Conclusion

Current pressures on the UK’s NHS have resulted in access to health care being challenging to almost all who use it. For those living on low incomes these challenges are exacerbated in many ways, as demonstrated by this research. Our findings show that despite NHS services being free at the point of delivery, low income can be a barrier to accessing health care. Poverty restricts access to health care for children and perpetuates inequities in health, education and life opportunities. These barriers are not inevitable. The findings of this research have informed CNE’s approach to working with health care providers to help them reduce the impacts poverty has on health care access using their ‘Poverty Proofing© Health audit approach’. The development of a ‘Common Themes Framework’ ensures audits across different settings are consistent, replicable, systematic and intelligence led.

## Data Availability

Due to the sensitive nature of the questions asked in this study and personal information shared, as well as our ethics approval, research participants were assured raw data would remain confidential and would not be shared.

## Acknowledgements

The authors wish to acknowledge the parent and VCSE participants who contributed to this study and the VCSE organisations who helped us engage with families. We would also like to thank Lorna Nicoll from CNE for proofreading this paper.

